# Secondary Analysis of Treatment Effect Heterogeneity of priovi in Borderline Personality Disorder

**DOI:** 10.64898/2026.11.09.26367607

**Authors:** Antje Riepenhausen, Gitta A. Jacob, Thomas Probst, Jan Philipp Klein

## Abstract

The digital therapeutic priovi has small average effects for borderline personality disorder (BPD). In this preregistered secondary analysis, we aimed to explore the heterogeneity and potential moderators of these effects. Adults received priovi plus treatment as usual or treatment as usual alone. At the 3-month primary endpoint (N = 528), model-based recursive partitioning showed heterogeneous effects on Borderline Symptom List-23 (BSL-23) change (*p* = 0.022) and on clinically relevant response (*p* = 0.007). Benefit was larger at higher baseline symptom load: a representative tree split on Patient Health Questionnaire-9 at a score of 23, and the higher-score subgroup had roughly twice the BSL-23 reduction (conditional average treatment effects −0.40 versus −0.19). Baseline BSL-23, depressive symptoms, and recent life-threatening events ranked as the leading moderators. Quartile calibration of predicted CATEs was weak and non-monotonic. The moderators are hypothesis-generating and should not guide patient selection without external validation.

## Introduction

Borderline personality disorder (BPD) is a common and burdensome condition associated with pronounced emotional instability, interpersonal difficulties, self-harm and elevated suicide risk, as well as substantial functional impairment and high health-care costs ^1,2^. Evidence-based psychotherapies exist but are resource-intensive and not universally accessible, motivating scalable digital interventions to bridge treatment gaps. priovi is a self-guided digital therapeutic based on schema-therapy principles, designed as an add-on to routine care. In the EPADIP-BPD randomized controlled trial, adding priovi to treatment as usual (TAU) produced a small average reduction in borderline symptom severity relative to TAU alone at the primary 3-month endpoint ^3^. As is typical for complex psychosocial and digital interventions, this average may obscure meaningful variation in individual benefit.

Understanding for whom a digital intervention works is central to precision mental health care. The central question in precision health care is “what works best for whom”. Addressing this question is relevant for patients, therapists, and the society. For an individual patient, it is important to know whether she:he with her:his individual characteristics will benefit from a digital intervention or not. If no benefit is expected, patients can save time and resources by skipping digital interventions without expected benefit for them and select and engage in digital interventions with a high probability of treatment success. Therapists also want to know if their individual patients will have a benefit from a digital intervention and whether they should recommend or prescribe a digital intervention to their individual patients. In societies with public reimbursement for empirically evaluated digital interventions, as for example in Germany for “DiGAs”, it is also of high financial relevance to know which patients with which characteristics benefit from a DiGA or not. Identifying the specific patient characteristics for which an intervention works most is challenging as there are potentially endless characteristics of patients that might be relevant to consider. Most of the past research, therefore, selected clinically relevant variables that might statistically moderate the effect of an intervention on the outcome to identify subgroups of patients with more or less treatment success ^4,5^. Yet, traditional one-moderator-at-a-time subgroup analyses are underpowered, prone to false positives under multiplicity, and unable to capture combinations of characteristics that jointly shape benefit. Model-based recursive partitioning (MOB-RP) and its ensemble extension, the model-based random forest, address these limitations by pairing a parametric treatment-effect model with an algorithmic search for regions of the moderator space in which that parameter shifts ^6,7^, producing interpretable, individualized estimates while guarding against overfitting.

In this preregistered secondary analysis of EPADIP-BPD, we applied MOB-RP to identify baseline characteristics that moderate the benefit of priovi on (i) change in BSL-23 and (ii) the probability of a clinically relevant response at the primary 3-month follow-up. We asked: (1) whether the treatment effect is heterogeneous; (2) which of seven prespecified candidate moderators are most influential; and (3) whether higher baseline psychopathology is associated with larger absolute benefit. Analyses at 6 and 12 months were prespecified as exploratory.

## Results

### Sample characteristics

The confirmatory T1 analysis sample comprised 528 participants with an observed BSL-23 outcome at three months (263 control, 265 intervention). Baseline characteristics by randomization arm are summarized in Table 1.

**Table 1.** Baseline characteristics of the T1 confirmatory analysis sample, by randomization arm.

| Characteristic | Control (TAU only; n = 263) | Intervention (priovi + TAU; n = 265) | Overall (n = 528) |
|---|---|---|---|
| Baseline BSL-23, mean (SD) | 2.3 (0.6) | 2.3 (0.7) | 2.3 (0.6) |
| Baseline PHQ-9, mean (SD) | 17.3 (4.6) | 17.2 (4.8) | 17.3 (4.7) |
| Baseline GAD-7, mean (SD) | 14.9 (3.8) | 14.7 (3.8) | 14.8 (3.8) |
| Suicide attempts, 3 months, mean (SD) | 0.1 (0.4) | 0.1 (0.7) | 0.1 (0.6) |
| Life-threatening events, 3 months, mean (SD) | 1.5 (5.5) | 1.3 (3.0) | 1.4 (4.4) |
| Psychotherapy at baseline, n (%) | 117 (44.5%) | 121 (45.7%) | 238 (45.1%) |
| Medication at baseline, n (%) | 126 (47.9%) | 126 (47.5%) | 252 (47.7%) |

*Abbreviations: TAU, treatment as usual; BSL-23, Borderline Symptom List-23; PHQ-9, Patient Health Questionnaire-9; GAD-7, 7-item Generalized Anxiety Disorder scale; SD, standard deviation. Group is coded 0 = control (TAU only) and 1 = intervention (priovi + TAU). BSL-23 is the mean item score (range 0-4). The table describes the primary (T1) complete-case analysis sample; baseline values are identical across follow-up samples up to the participants excluded by outcome missingness.*

### Differential effect of priovi on borderline symptom severity at T1 (confirmatory)

The confirmatory analysis of change in BSL-23 at T1 was based on N = 528 participants. The omnibus test rejected the global null of a homogeneous treatment effect (p = 0.022), supporting the primary hypothesis of treatment-effect heterogeneity.

By conditional permutation importance, the most influential moderators were Baseline BSL-23, Baseline PHQ-9, and Life-threatening events (3 months) (Table 2), identifying baseline psychopathology as the dominant class of moderators.

**Table 2.** Conditional permutation variable importance (BSL-23 change at T1).

| Moderator | Importance | 95% CI | Root-split freq. |
|---|---|---|---|
| Baseline BSL-23 | 4479.481 | [3636.741, 15020.252] | 5% |
| Baseline PHQ-9 | 2616.903 | [1855.601, 8023.515] | 70% |
| Life-threatening events (3 months) | 1242.462 | [1028.786, 5312.894] | 10% |
| Medication at baseline | 1079.178 | [971.092, 4625.204] | 9% |
| Baseline GAD-7 | 924.734 | [1035.058, 4295.707] | 4% |
| Psychotherapy at baseline | 667.503 | [652.485, 3659.833] | 2% |
| Suicide attempts (3 months) | 0.000 | [0.000, 0.000] | 0% |

*Abbreviations: BSL-23, Borderline Symptom List-23; PHQ-9, Patient Health Questionnaire-9; GAD-7, 7-item Generalized Anxiety Disorder scale; CI, confidence interval. Importance is the conditional permutation variable importance (unitless; larger values indicate a more influential moderator), with 95% bootstrap CIs. Root-split frequency is the percentage of bootstrap forest trees in which the moderator defined the first (root) split.*

Consistent with this, the representative tree split on Baseline PHQ-9 at a cut-point of 23 (Figure 1). The subgroup with the larger benefit comprised participants with higher baseline PHQ-9 (terminal node 3, n = 60) and had a CATE of -0.40 BSL-23 points, versus -0.19 points in the smaller-benefit subgroup (node 2, n = 468); more negative values denote a greater reduction in symptom severity favoring priovi. The 95% bootstrap CIs of both subgroup CATEs excluded zero (Table 3), but they overlapped substantially and the higher-symptom subgroup was small, so the difference between subgroups is not firmly established. This split was the most stable feature of the analysis: Baseline PHQ-9 was selected as the root moderator in 70% of bootstrap trees. Notably, the single sharpest split point was thus defined by baseline depressive symptoms rather than by baseline BSL-23 itself, even though baseline BSL-23 ranked among the most important moderators overall.

**Figure 1.**
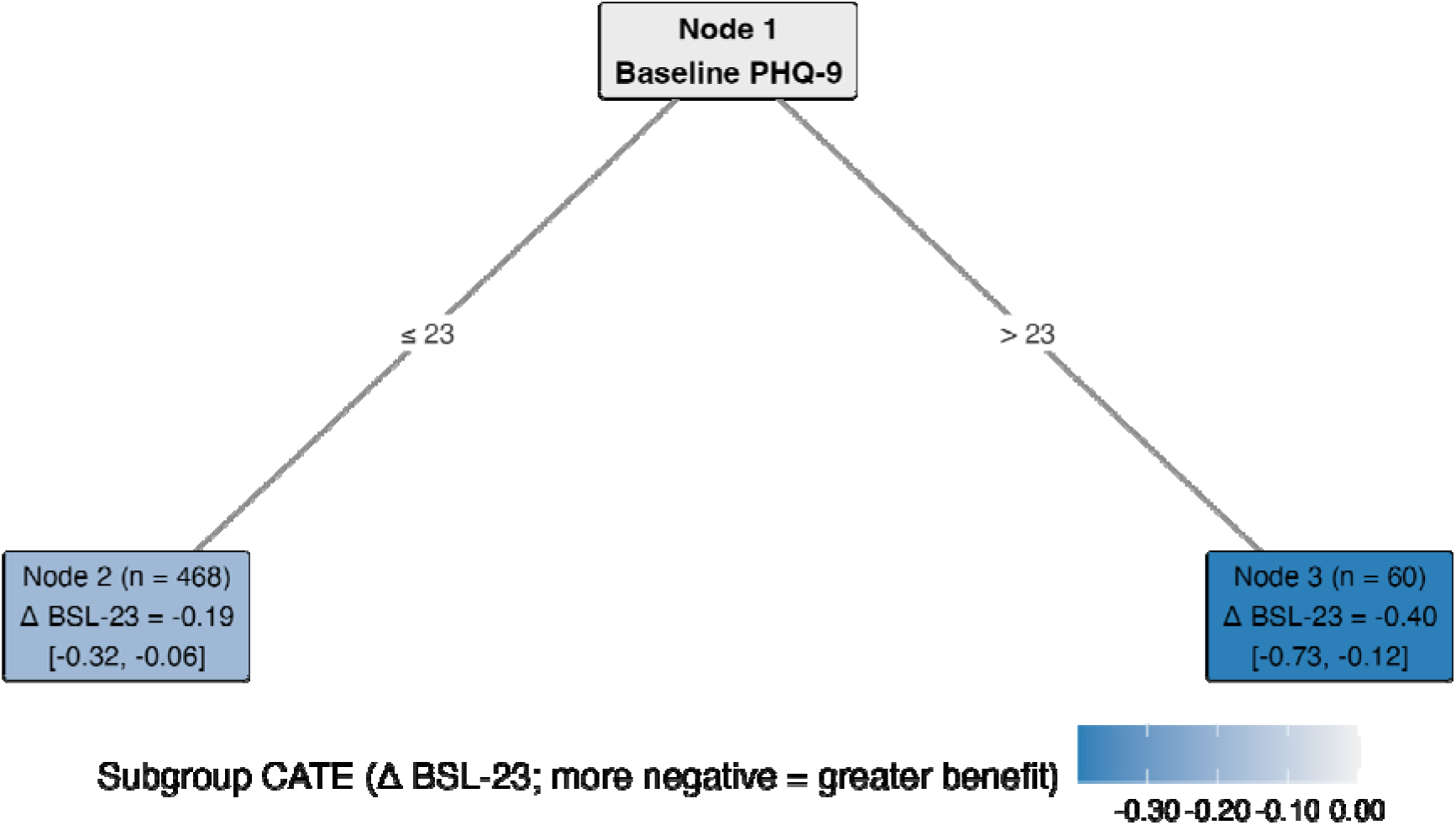
Representative model-based tree for change in BSL-23 at T1. Internal nodes show the splitting baseline variable; edges give the split condition. Each terminal node reports its size and adjusted subgroup CATE with the 95% bootstrap confidence interval, and the leaf fill encodes the CATE (more negative = greater reduction in symptom severity favoring priovi).

**Table 3.** Subgroup conditional average treatment effects (BSL-23 change at T1).

| Subgroup | Estimated CATE | 95% CI |
|---|---|---|
| Terminal node 2 | -0.19 | [-0.32, -0.06] |
| Subgroup | Estimated CATE | 95% CI |
| Terminal node 3 | -0.40 | [-0.73, -0.12] |

*Abbreviations: CATE, conditional average treatment effect; CI, confidence interval. The estimated CATE is the subgroup-specific mean difference in BSL-23 change (priovi vs control); more negative values indicate greater benefit. The 95% CIs are bootstrap percentile intervals.*

The directional secondary hypothesis (that higher baseline psychopathology is associated with larger absolute benefit) was supported descriptively, as the subgroup defined by higher baseline depressive symptoms showed the larger reduction in BSL-23. Calibration across quartiles of the predicted CATE was weak and non-monotonic (Figure 2; *r* = -0.65): predicted and observed subgroup effects were inversely associated across quartiles, indicating poor calibration of the prediction model despite detectable group-level heterogeneity.

**Figure 2.**
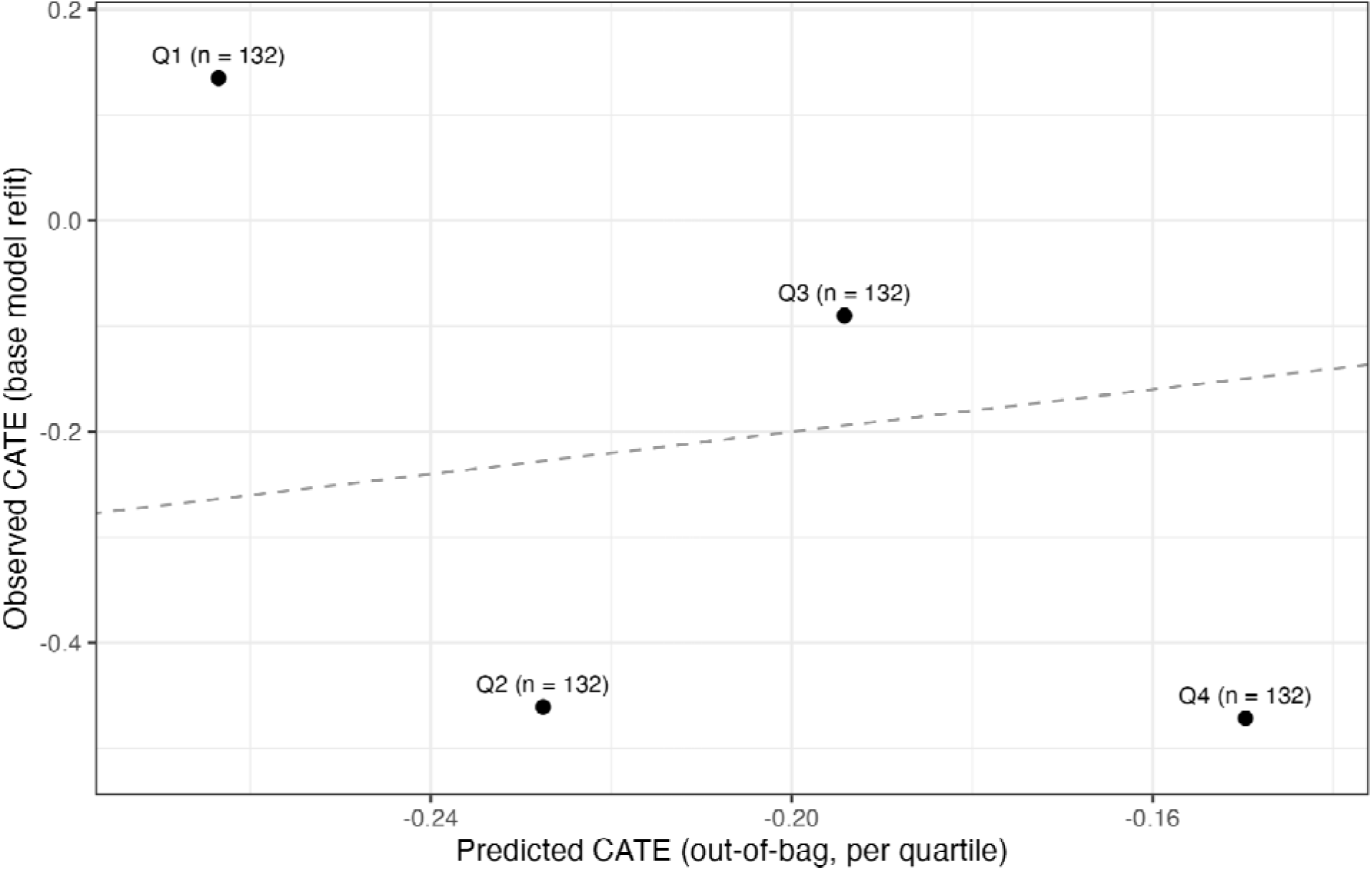
Calibration of the predicted treatment effect: mean predicted versus observed subgroup CATE across quartiles of the out-of-bag predicted CATE (BSL-23 at T1). Points are labeled by quartile and subgroup size (n); the dashed line marks perfect calibration.

### Differential effect of priovi on treatment response at T1

Because the BSL-23 omnibus test at T1 was significant (*p* = 0.022), the responder analysis is interpreted confirmatorily.

The responder analysis was based on N = 528 participants (41% responders under control vs. 57% under priovi). The omnibus heterogeneity test yielded *p* = 0.007. The most influential moderator by conditional permutation importance was Baseline BSL-23, followed by Medication at baseline and Psychotherapy at baseline (Table 4).

**Table 4.** Conditional permutation variable importance (treatment response at T1).

| Moderator | Importance | 95% CI | Root-split freq. |
|---|---|---|---|
| Baseline BSL-23 | 6.357 | [3.097, 17.359] | 66% |
| Medication at baseline | 2.139 | [1.049, 9.228] | 14% |
| Psychotherapy at baseline | 2.126 | [0.739, 9.023] | 15% |
| Baseline GAD-7 | 1.806 | [0.371, 8.175] | 4% |
| Life-threatening events (3 months) | 1.269 | [0.304, 6.573] | 0% |
| Baseline PHQ-9 | 1.096 | [0.778, 7.643] | 0% |
| Suicide attempts (3 months) | 0.000 | [0.000, 0.000] | 0% |

*Abbreviations: BSL-23, Borderline Symptom List-23; PHQ-9, Patient Health Questionnaire-9; GAD-7, 7-item Generalized Anxiety Disorder scale; CI, confidence interval. Importance is the conditional permutation variable importance (unitless; larger values indicate a more influential moderator), with 95% bootstrap CIs. Root-split frequency is the percentage of bootstrap forest trees in which the moderator defined the first (root) split.*

The representative tree split on Baseline BSL-23 at a cut-point of 1.83 (Figure 3), and Baseline BSL-23 was selected at the root in 66% of bootstrap trees, making it a highly stable moderator. Notably, however, the estimated risk difference in favor of priovi was virtually identical across the resulting subgroups (approximately +0.15 in both; Table 5), so the split separated patients’ baseline probability of responding rather than their benefit from priovi; the corresponding subgroup confidence intervals overlapped almost completely.

**Figure 3.**
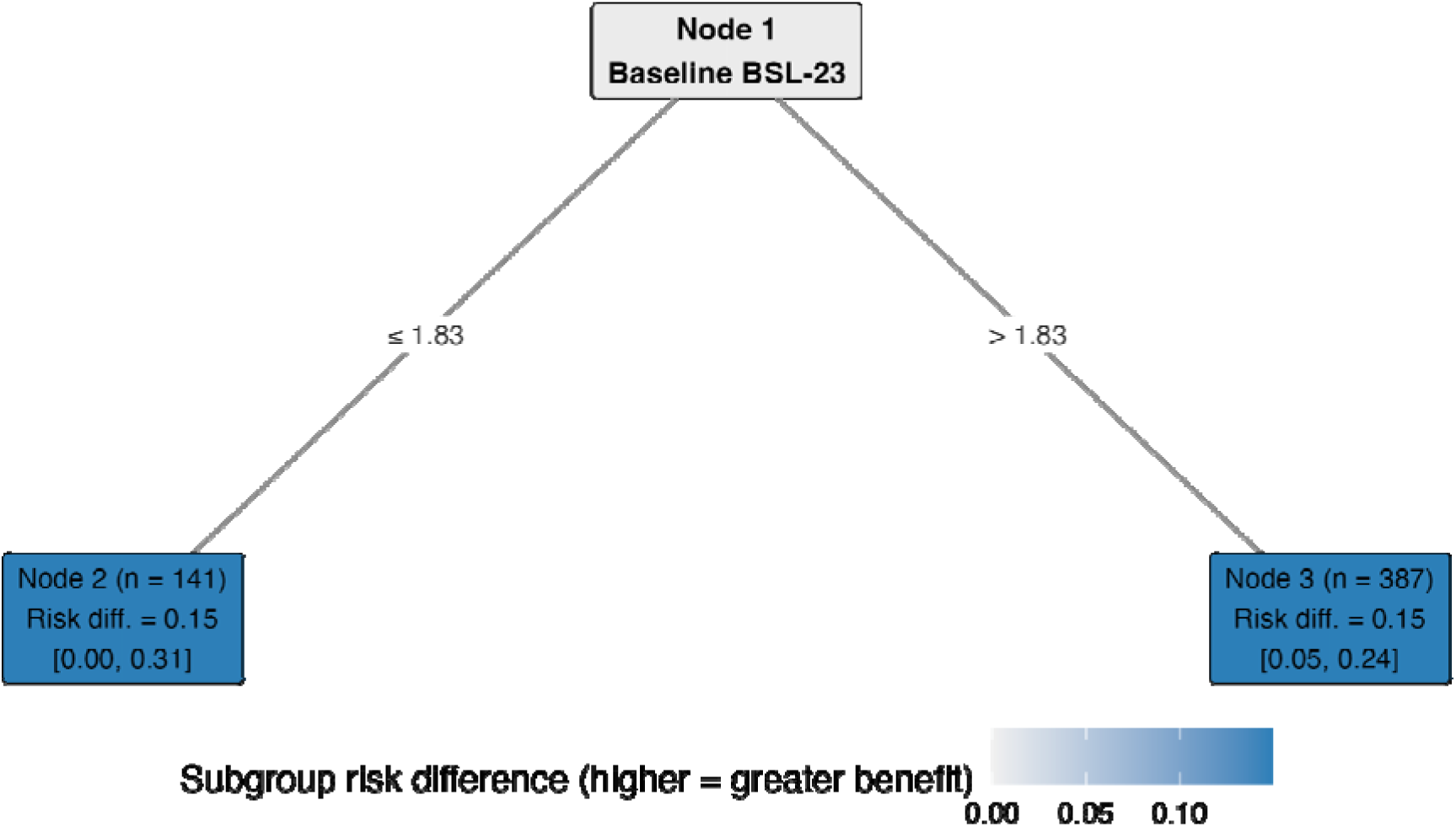
Representative model-based tree for treatment response at T1. Internal nodes show the splitting baseline variable; edges give the split condition. Each terminal node reports its size and subgroup risk difference with the 95% bootstrap confidence interval, and the leaf fill encodes the effect (higher = greater probability of response favoring priovi).

**Table 5.** Subgroup conditional average treatment effects (treatment response at T1, risk difference).

| Subgroup | Estimated<br>CATE | 95% CI |
|---|---|---|
| Terminal node 2 | 0.15 | [0.00, 0.31] |
| Terminal node 3 | 0.15 | [0.05, 0.24] |

*Abbreviations: CATE, conditional average treatment effect; CI, confidence interval. For the binary responder outcome the estimated CATE is a risk difference (probability of response under priovi minus control); higher values indicate greater benefit. The 95% CIs are bootstrap percentile intervals.*

Individual-level discrimination was limited: the concordance-for-benefit statistic was 0.47, close to the 0.5 chance level, indicating that the model did not reliably rank patients by their benefit from priovi.

### Exploratory analyses: stability across time (T2 and T3)

#### BSL-23 change at T2 (6 months) (N = 502)

The omnibus heterogeneity test yielded *p* = 0.009. No split survived the representative tree’s Bonferroni threshold, indicating that any heterogeneity was diffuse rather than attributable to a single sharp subgroup; the overall CATE was -0.14 BSL-23 points (95% CI excluding zero). Even so, Baseline PHQ-9 was the most frequent root moderator across bootstrap trees (33%). Quartile calibration of the predicted CATE was moderate (*r* = 0.71).

#### BSL-23 change at T3 (12 months) (N = 451)

The omnibus heterogeneity test yielded *p* = 0.004. The representative tree split on Medication at baseline (presence vs. absence): the more favorable estimate was seen in participants with medication at baseline = no (node 2, n = 235), with a CATE of 0.02 BSL-23 points versus 0.05 in the other subgroup (node 3, n = 216), and the 95% CI excluded zero for neither subgroup. Medication at baseline was the most frequent root moderator (61% of bootstrap trees). Quartile calibration of the predicted CATE was weak (*r* = -0.03).

#### Treatment response at T2 (6 months) (N = 502)

The omnibus heterogeneity test yielded p < .001. The representative tree split on Baseline BSL-23 (cut-point 1.78): the more favorable estimate was seen in participants with higher baseline BSL-23 (node 3, n = 377), with a CATE of 0.12 risk-difference points versus 0.02 in the other subgroup (node 2, n = 125), and the 95% CI excluded zero for node 3. Baseline BSL-23 was the most frequent root moderator (76% of bootstrap trees). The concordance-for-benefit statistic was 0.50.

#### Treatment response at T3 (12 months) (N = 451)

The omnibus heterogeneity test yielded *p* = 0.031. No split survived the representative tree’s Bonferroni threshold, indicating that any heterogeneity was diffuse rather than attributable to a single sharp subgroup; the overall CATE was 0.01 risk-difference points (95% CI including zero). Even so, Baseline BSL-23 was the most frequent root moderator across bootstrap trees (83%). The concordance-for-benefit statistic was 0.51.

For treatment response, baseline BSL-23 was the most stable moderator across all three time points, selected at the root of bootstrapped trees in 66%, 76% and 83% of resamples at T1, T2 and T3, respectively. As at T1, baseline symptom severity (baseline BSL-23 and PHQ-9) dominated the conditional importance rankings for the continuous outcome, and where an interpretable subgroup emerged the larger benefit again fell to patients with higher baseline severity, consistent with the directional secondary hypothesis. The exploratory analyses also revealed limits to this pattern: at some time points the omnibus test was significant while no single split survived the representative tree’s Bonferroni threshold; subgroup estimates were imprecise with widely overlapping confidence intervals; and individual-level validation remained weak (concordance-for-benefit near 0.50). By the 12-month follow-up the subgroup treatment effects were close to zero with confidence intervals including zero, in line with an intervention benefit that attenuates over time. These time points were prespecified as exploratory and are not interpreted confirmatorily.

Conditional and unconditional (which does not account for correlations among the candidate moderators) variable importance are compared for every analysis in the Supplementary Information (Supplementary Tables 1 - 6). Partial dependence plots for both outcomes and all time points can be found in the Supplementary Information (Supplementary Fig. 1 - 6).

## Discussion

Adding priovi to treatment as usual produced a heterogeneous rather than a uniform benefit at the three-month endpoint. For borderline symptom severity, the global test of heterogeneity was significant (*p* = 0.022), and both the representative tree and the bootstrap root-split frequency identified baseline depressive symptoms as the most stable moderator, whereas conditional variable importance ranked baseline BSL-23 highest; both findings point to baseline symptom severity as the central axis of moderation. Patients with a higher baseline symptom load showed the larger absolute reduction in BSL-23, consistent with our directional hypothesis. For clinically relevant response the heterogeneity test was likewise significant (*p* = 0.007) and baseline BSL-23 was the dominant, highly stable moderator, but the resulting subgroups differed mainly in their baseline probability of responding rather than in the absolute benefit derived from priovi. Across both outcomes, predictive performance was weak: quartile calibration for BSL-23 change was non-monotonic and the concordance-for-benefit for response was close to chance, indicating that, although moderation is detectable at the group level, the present model is not ready to guide individual-level treatment decisions. Exploratory analyses at 6 and 12 months were broadly consistent in identifying baseline symptom severity as the recurring moderator, most stably for treatment response, but the subgroup structure became less clearly defined and the treatment benefit attenuated toward the 12-month follow-up, underscoring that these moderators are hypothesis-generating and require prospective external validation.

In the main EPADIP-BPD trial, adding priovi to treatment as usual produced only a small average reduction in borderline symptom severity at the three-month endpoint ^3^. This modest average effect is consistent with the broader treatment literature for BPD, in which even established face-to-face psychotherapies such as dialectical behavior therapy and psychodynamic approaches produce only small to moderate effects that are frequently unstable at follow-up ^8,9^. It also aligns with the emerging evidence on digital interventions for BPD, where benefits have been shown for specific targets such as suicidal ideation but not consistently for overall symptom severity ^10,11^. Our analysis qualifies this average: the benefit was not uniform but was concentrated among patients with higher baseline symptom severity, in whom the estimated reduction in BSL-23 was approximately twice as large. A pattern in which more severely affected patients derive larger absolute benefit, having more room for improvement, has been described for other psychological and digital interventions ^12^. In particular, an individual-patient-data network meta-analysis of internet-based cognitive behavioral therapy for depression identified baseline severity as the most important modifier of relative efficacy, with larger benefits at higher baseline severity ^13^, closely paralleling the severity gradient we observed for priovi. Importantly, in the binary responder analysis higher baseline severity was also associated with a higher baseline probability of response, so part of the apparent moderation reflects differences in baseline prognosis rather than in the treatment benefit itself, a distinction between prognostic (risk) modeling and effect modification ^14^ that is central to predictive approaches to treatment-effect heterogeneity ^15,16^. Methodologically, our approach applies established model-based recursive partitioning and model-based forest methods for subgroup and individualized treatment-effect estimation ^6,7^ together with conditional permutation importance ^17^. Finally, the weak out-of-bag calibration and near-chance concordance-for-benefit we observed are consistent with broader concerns that reliable individual-level treatment-benefit prediction is difficult to achieve in single trials of moderate size ^18^.

Our analysis has several strengths. First, it followed a preregistered protocol, which fixed the outcomes, candidate moderators, and analytic strategy in advance and thereby kept the confirmatory three-month analysis clearly separate from the exploratory follow-up analyses, reducing the risk of data-driven or selective reporting. Second, rather than testing one candidate moderator at a time, we used a data-adaptive approach (model-based recursive partitioning and model-based forests) that weighs many baseline characteristics jointly and lets the data determine which of them, and in what combination, modify the treatment effect, while built-in complexity control guards against overfitting. Third, we assessed predictive performance honestly: calibration and the concordance-for-benefit were estimated on out-of-bag observations that had not contributed to the trees generating a given prediction, so they reflect genuine out-of-sample behavior rather than in-sample optimism. Fourth, we quantified uncertainty with a distribution-free bootstrap, obtaining subgroup confidence intervals that do not depend on parametric assumptions about the shape of the treatment-effect distribution.

These strengths are balanced by several limitations that temper our conclusions. Because the analyses were complete-case in the presence of differential dropout, their validity rests on the assumption that missingness is conditionally ignorable, that is, missing at random given the observed baseline variables and randomization arm; if unmeasured factors jointly drive both dropout and outcome, the estimates could be biased in ways the present design cannot rule out. Statistical power adds a further constraint, as the study was adequately powered only for moderate heterogeneity (standardized subgroup differences of about d ≥ 0.35) and underpowered for smaller but potentially meaningful differences in benefit, so the absence of a given moderator signal cannot be read as firm evidence that no such moderation exists.

A second set of caveats concerns how the severity signal should be interpreted. The candidate moderators were baseline characteristics that were observed rather than manipulated, and although randomization renders the average treatment effect unconfounded, the association between a moderator and the magnitude of benefit is itself observational and may be confounded by unmeasured correlates of baseline severity. This is compounded by the construction of the responder outcome, which combines a reliable-change criterion with a shift to a less severe severity grade and uses baseline BSL-23 both as the reference for change and as a candidate moderator; together these features can make baseline severity mechanically influential and should be borne in mind when interpreting its prominence. Precision was a further concern: the subgroup with the largest benefit at T1 was small (n = 60) and its confidence interval wide, while calibration and the concordance-for-benefit indicated poor individual-level discrimination, so the moderation is best regarded as hypothesis-generating rather than clinically actionable. The same caution applies to the later time points, where analyses beyond the primary endpoint were exploratory and multiple and the attenuation of effects by twelve months coincided with degenerate representative trees, limiting their interpretation. One prespecified moderator, suicide attempts in the previous three months, was too rare (reported by only 6.4% of participants) to yield an admissible split in any tree, so its zero variable importance reflects a lack of statistical resolution for a low-prevalence variable rather than a moderator that was evaluated and excluded. Finally, because the findings derive from a single trial with differential dropout and were not externally validated, their generalizability remains uncertain ^16,19^.

priovi is a low-threshold, scalable, and low-risk digital add-on to standard care, and our results speak to how confidently it can be offered across the range of patients seen in practice. Baseline symptom severity was the most consistent signal across outcomes and time points, and patients with a higher baseline symptom load showed the largest absolute reduction in BSL-23. Because more severely affected patients often carry the greatest unmet need and can be harder to reach with resource-intensive care, it is encouraging that a self-guided digital option appears to benefit them at least as much as others. This lends additional support to offering priovi broadly as a complement to standard treatment, with particular promise for patients presenting with greater baseline severity. These implications remain provisional: individual-level calibration was weak, concordance-for-benefit was close to chance, and the subgroup benefit attenuated by twelve months, so the results do not yet support tailoring treatment to individual patients. Baseline severity, like any other candidate moderator, would need to be confirmed prospectively and validated in independent samples before it could formally inform clinical practice ^20,21^.

In this preregistered secondary analysis, the three-month benefit of priovi was heterogeneous and most pronounced in patients with higher baseline symptom severity, yet validation of the predicted treatment effects was weak and the effect attenuated by twelve months. Baseline severity is thus a promising but not yet actionable moderator. More broadly, this first preregistered heterogeneity analysis of a digital therapeutic for BPD shows that group-level moderation can be detected while reliable prediction of treatment benefit remains out of reach in a single trial of moderate size. Realizing the promise of personalized digital care will require prospective external validation in larger, ideally multi-site samples, and evaluation of the clinical utility of acting on such predictions, before baseline severity or any other moderator can guide patient selection.

## Methods

### Study design and participants

This is a preregistered secondary analysis of the EPADIP-BPD trial; its full design, procedures and primary results are reported in the main publication ^3^. In brief, that trial enrolled adults whose BPD diagnosis was confirmed with the SCID-5-PD and who showed at least moderate symptom severity (a BSL-23 mean item score ≥ 1.07), while excluding people with a comorbid substance use or psychotic disorder. Using a pragmatic, rater-blinded, two-arm parallel-group design, 580 participants were randomized in Germany, 302 to priovi added to TAU and 278 to TAU alone. Each participant was assessed before randomization (T0) and again 3, 6 and 12 months later (T1, T2 and T3). T1 was the trial’s primary endpoint and is the primary, confirmatory time point here; T2 and T3 were analyzed as exploratory sensitivity analyses. Because this secondary analysis was conducted on a complete-case basis (see below), the analyzed sample size varies across follow-ups, reflecting the number of participants with an observed outcome at each time point rather than the full randomized sample. The EPADIP-BPD trial was registered in the German Clinical Trials Register (DRKS00028888; https://drks.de/search/de/trial/DRKS00028888; registration date: 27 April 2022). It was reviewed and approved by the ethics committee of the University of Lübeck (reference number 22-012), and all participants gave informed written consent to participate ^3^. The original approval covers the present secondary analysis of the trial data. This secondary analysis was preregistered on the Open Science Framework (https://osf.io/sz72e/).

### Treatment condition

The randomization arm (group) is the treatment variable of primary interest, coded as a binary factor with levels 0 (control; TAU only) and 1 (intervention; priovi added to TAU). Because all candidate moderators were measured at baseline, they are independent of the arm by design.

## Measures

### Outcomes

The BSL-23 was scored as the mean item score (the sum of the 23 items divided by 23; range 0-4), and all BSL-23 quantities below are reported on this scale. The continuous outcome was the change in the BSL-23 ^22^ mean item score from baseline to the respective follow-up (T0-to-T1 for the confirmatory analysis). The binary outcome was clinically relevant response, defined as meeting both the reliable-change-index criterion ^23^ and showing a change in the BSL-23 score toward a less severe severity grade relative to baseline ^24^ (1 = responder, 0 = non-responder).

### Baseline covariate

For the BSL-23 outcome, the baseline BSL-23 mean item score entered the base model as a covariate to adjust for regression to the mean.

### Candidate moderators

Seven prespecified baseline variables were supplied as candidate moderators: (i) baseline BSL-23, (ii) baseline PHQ-9 ^25^, (iii) baseline GAD-7 ^26^, (iv) number of suicide attempts in the three months before baseline, (v) number of life-threatening events in the three months before baseline, (vi) psychotherapy status at baseline (1/0), and (vii) medication status at baseline (intake of sedatives or antidepressants; 1/0). Medication status was derived from recorded Anatomical Therapeutic Chemical (ATC) codes as any regular or as-needed intake of an agent classified under N06A (antidepressants) or N05C (hypnotics/sedatives). Baseline BSL-23 enters in two roles (as a covariate in the BSL-23 base model and as a candidate moderator) so that partitioning can detect moderation by baseline severity; splitting responds to instability in the treatment-effect parameter itself (detected via the score function), not to its average level.

### Missing data and analysis samples

We used complete-case analysis throughout: for each follow-up we included only participants whose outcome at that time point was observed, and we did not impute missing outcomes. Because complete baseline data were a prerequisite for randomization, inclusion is governed solely by outcome availability. Dropout differed between arms (higher under the intervention).

### Statistical analysis

Following our preregistered plan, we used model-based recursive partitioning (MOB-RP) and model-based random forests to identify combinations of baseline characteristics that jointly modify the effect of priovi ^6,7^. Both outcomes were analyzed separately at the primary 3-month endpoint (T1). The same procedure was repeated at 6 and 12 months (T2 and T3) as exploratory analyses. The primary analyses were complete-case: participants without an observed follow-up BSL-23 at the relevant time point were excluded. As a robustness check against differential dropout, we repeated the two T1 analyses after a single stochastic imputation of follow-up BSL-23 (predictive mean matching, fitted separately within each randomization arm, one completed dataset, seed 1234); the binary responder was derived from the imputed BSL-23 using the same definition as in the primary analysis. Full single-imputation results confirm the complete-case analysis and are reported in the Supplementary Information (Supplementary Note 1 and Supplementary Tables 7 – 11). The complete-case analysis remains the primary analysis.

### Base models

For each outcome we specified a simple model whose treatment-effect parameter was the target of partitioning. For the continuous outcome this was a baseline-adjusted (ANCOVA-type) linear model of the change in BSL-23, with baseline BSL-23 as a covariate. For the binary responder outcome it was a logistic model estimated with bias reduction, which keeps the treatment-effect estimate finite even in small subgroups while remaining virtually identical to maximum likelihood otherwise. Model-based recursive partitioning then searched the seven prespecified baseline moderators (baseline BSL-23, PHQ-9, GAD-7, suicide attempts, life-threatening events, current psychotherapy, and current medication) for cut-points across which the model parameters changed. Because this instability is assessed over the whole score vector of the base model, a split can reflect a difference in the size of the treatment benefit, a difference in baseline prognosis, or both.

### Forest, omnibus test, and variable importance

To estimate this heterogeneity robustly, we aggregated 500 such trees into a model-based random forest. Each tree was fitted to a group-stratified subsample of 63.2% of observations drawn without replacement; at each split, three candidate moderators were considered at random, with a minimum of 50 observations per terminal node. We tested the global null hypothesis that the treatment effect does not vary with any baseline characteristic using a parametric-bootstrap permutation test with 1000 replicates. We ranked the moderators by conditional permutation variable importance ^17^, which evaluates each moderator within strata defined by the remaining moderators and therefore reflects its contribution conditional on the others. Unconditional permutation importance, which permutes each moderator independently and therefore reflects its marginal association, is reported in the Supplementary Information (Supplementary Tables 1 – 6).

### Model assumptions and inference

The omnibus heterogeneity test used a parametric-bootstrap permutation procedure (B = 1000) based on score contributions from the base model and therefore does not depend on residual normality of the linear BSL-23 model. For the binary outcome, bias-reduced logistic regression was used to avoid infinite estimates in small subgroups.

### Representative tree, subgroup effects, and partial dependence

For interpretability we fitted a single representative tree with the same base model and the same seven moderators, Bonferroni-corrected split tests across those moderators (minimum criterion 0.95), and a minimum of 50 observations per terminal node. For each terminal subgroup we report the conditional average treatment effect (CATE): the group coefficient for the continuous outcome and a g-computation risk difference for the binary outcome. For the three most important moderators we also visualized how the predicted effect varied across the moderator, averaging over the observed distribution of the remaining moderators (partial-dependence profiles; Supplementary Fig. 1 - 6).

### Uncertainty, calibration, and discrimination

Uncertainty for the importance scores and the subgroup CATEs was obtained from a stratified nonparametric bootstrap (1000 resamples with replacement within arm). The tree structure was fixed on the original data and re-evaluated in each resample; 95% confidence intervals are the 2.5th and 97.5th percentiles of the bootstrap distribution. We also recorded how often each moderator defined the first split of a tree refitted in each resample. For the continuous outcome we assessed calibration by grouping out-of-bag predicted CATEs into quartiles and comparing them with the observed treatment effect in each quartile. For the binary outcome we computed the concordance-for-benefit statistic from out-of-bag predictions ^15^. Both metrics used the same stratified bootstrap.

### Multiplicity

All confirmatory tests were two-sided at α = .05. The two outcomes were tested hierarchically, BSL-23 first, with the responder analysis interpreted confirmatorily only if the BSL-23 omnibus test was significant and exploratorily otherwise. Only the T1 analyses were confirmatory. Subgroups whose 95% CI for the CATE excluded zero were interpreted as evidence of meaningful heterogeneity; the directional secondary hypothesis was evaluated descriptively.

### Statistics and reproducibility

Analyses used R (v4.6.1) ^27^ with model4you (v0.9-9) ^28^, partykit (v1.2-27) ^29^, brglm2 (v1.1.0) ^30^ and ggplot2 (v4.0.3) ^31^; forest fitting and the bootstrap were parallelized. A fixed seed (set.seed(1234)) was set before analysis and propagated to the bootstrap. The code used for analysis can be found at https://osf.io/ker2t/.

### Patient and public involvement

Patient and public involvement. Patients and the public were not involved in the design, conduct, analysis, or reporting of this secondary analysis. In the parent EPADIP-BPD trial, individuals with BPD were involved in the development of priovi, but not in the design of the trial itself ^3^.

### Use of large language models

A large language model accessed via the Cursor coding assistant was used to polish the language of the manuscript and to assist with manuscript formatting for journal style. The authors reviewed and edited all text and take full responsibility for the content.

### Deviations from the preregistration

Two participants (both intervention group) had missing values on one or more baseline candidate moderators and were excluded from all analyses; the confirmatory T1 sample therefore comprised 528 participants rather than the preregistered 530. The full pipeline (500 trees; pmtest B = 1000; bootstrap B = 1000) was run as specified.

## Data availability

The dataset analyzed here is not publicly available. Individual participant data underlying the results can be shared with researchers who provide a methodologically sound proposal to J. P. Klein. Proposals can be submitted up to 36 months after publication of this Article.

## Code availability

The full analysis code, package versions and random seed, together with the preregistration (registered on the Open Science Framework on 7 July 2026), are available at https://osf.io/ker2t/.

## Acknowledgments

Open access funding provided by University of Vienna. The secondary analysis received no specific funding. The original EPADIP-BPD trial was funded by GAIA (Hamburg, Germany), which developed and operates priovi. We thank all participants of the EPADIP-BPD trial. Ethics approval for the trial, which covers the present secondary analysis, is described in the Methods.

## Author contributions

Conceptualization and methodology: AR, GAJ, JPK, and TP. Formal analysis, visualization, and writing – original draft: AR. Writing – review and editing: GAJ, TP, and JPK. All authors read and approved the final manuscript.

## Competing interests

priovi is developed and operated by GAIA (Hamburg, Germany), the funder of the original EPADIP-BPD trial. AR and GAJ are employees of GAIA; GAJ has additionally received payments for schema-therapy training from schema-therapy training institutes and royalties for books and DVDs on schema therapy and the treatment of BPD from Hogrefe and Beltz. JPK declared funding for clinical trials from the German Federal Ministry of Health and Servier; payments for presentations from GAIA, Oberberg, Servier, and Stillachhaus; consulting fees from All About Me, Ethypharm, and GAIA; payments for workshops and books from Beltz, Elsevier, Hogrefe, and Springer; and serving as vice-chair of the Digital Psychiatry section of the German Psychiatric Association. TP reports consulting fees from Stiftung Warentest and Health Study Club.

## Notes

### Clinical Trial

DRKS00028888

### Author Declarations

The ethics committee of the University of Lübeck gave ethical approval for this work (reference number 22-012).

