## Supplementary Material for "Secondary Analysis of Treatment Effect Heterogeneity of priovi in Borderline Personality Disorder"

### **S1. Conditional versus unconditional variable importance**

For each model-based random forest we quantified the influence of a candidate moderator by permuting its values and recording the resulting decrease in the forest’s objective function. Two variants were computed. **Unconditional** (marginal) importance permutes a moderator while ignoring the others; when moderators are correlated—as baseline symptom scales typically are—this can spread importance across all correlated variables and overstate the apparent influence of variables that merely proxy a true driver. **Conditional** permutation importance ^1^ permutes a moderator within strata defined by the remaining moderators, thereby isolating the information a moderator contributes over and above its correlates. Conditional importance is therefore the primary metric reported in the main text; the unconditional values are provided here for completeness and transparency.

The tables below list both metrics for all baseline candidate moderators, sorted by conditional importance, together with each moderator’s rank under the two metrics, a 95% confidence interval for the conditional metric, and the root-split frequency (the proportion of stratified bootstrap trees in which the moderator was selected at the root), all obtained from the same stratified nonparametric bootstrap. Because importance is measured on the scale of the model objective function, the magnitudes are comparable across moderators within a given analysis but not between analyses or between the continuous and binary outcomes.

**Supplementary Table 1. Variable importance for change in BSL-23 at T1 (confirmatory).**

| Moderator | Cond. imp. | Uncond. imp. | Rank (cond.) | Rank (uncond.) | Conditional 95% CI | Root-split freq. |
| --- | --- | --- | --- | --- | --- | --- |
| Baseline BSL-23 | 4479.481 | 4536.459 | 1 | 1 | [3636.741, 15020.252] | 5% |
| Baseline PHQ-9 | 2616.903 | 2686.882 | 2 | 2 | [1855.601, 8023.515] | 70% |
| Life-threatening events (3 months) | 1242.462 | 1269.344 | 3 | 3 | [1028.786, 5312.894] | 10% |
| Medication at baseline | 1079.178 | 1124.018 | 4 | 4 | [971.092, 4625.204] | 9% |
| Baseline GAD-7 | 924.734 | 899.941 | 5 | 5 | [1035.058, 4295.707] | 4% |
| Psychotherapy at baseline | 667.503 | 659.467 | 6 | 6 | [652.485, 3659.833] | 2% |
| Suicide attempts (3 months) | 0.000 | 0.000 | 7 | 7 | [0.000, 0.000] | 0% |

*N = 528. Importance is expressed on the scale of the model objective function (residual sum of squares (continuous BSL-23 model)); values are comparable across moderators within this analysis but not across analyses. Cond. = conditional. Uncond. = unconditional. Imp. = importance. Freq. = frequency.*

**Supplementary Table 2. Variable importance for treatment response at T1.**

| Moderator | Cond. imp. | Uncond. imp. | Rank (cond.) | Rank (uncond.) | Conditional 95% CI | Root-split freq. |
| --- | --- | --- | --- | --- | --- | --- |
| Baseline BSL-23 | 6.357 | 6.257 | 1 | 1 | [3.097, 17.359] | 66% |
| Medication at baseline | 2.139 | 2.087 | 2 | 3 | [1.049, 9.228] | 14% |
| Psychotherapy at baseline | 2.126 | 2.111 | 3 | 2 | [0.739, 9.023] | 15% |
| Baseline GAD-7 | 1.806 | 1.739 | 4 | 4 | [0.371, 8.175] | 4% |
| Life-threatening events (3 months) | 1.269 | 1.462 | 5 | 5 | [0.304, 6.573] | 0% |
| Baseline PHQ-9 | 1.096 | 1.059 | 6 | 6 | [0.778, 7.643] | 0% |
| Suicide attempts (3 months) | 0.000 | 0.000 | 7 | 7 | [0.000, 0.000] | 0% |

*N = 528. Importance is expressed on the scale of the model objective function (log-likelihood (binary responder model)); values are comparable across moderators within this analysis but not across analyses. Cond. = conditional. Uncond. = unconditional. Imp. = importance. Freq. = frequency.*

**Supplementary Table 3. Variable importance for change in BSL-23 at T2 (exploratory).**

| Moderator | Cond. imp. | Uncond. imp. | Rank (cond.) | Rank (uncond.) | Conditional 95% CI | Root-split freq. |
| --- | --- | --- | --- | --- | --- | --- |
| Baseline BSL-23 | 3058.058 | 3060.827 | 1 | 1 | [3382.085, 12924.145] | 6% |
| Baseline PHQ-9 | 2952.526 | 3006.432 | 2 | 2 | [2086.972, 8328.814] | 33% |
| Medication at baseline | 1811.297 | 1807.179 | 3 | 3 | [1148.796, 5569.896] | 29% |
| Life-threatening events (3 months) | 1271.210 | 1232.620 | 4 | 4 | [1042.577, 4744.883] | 24% |
| Psychotherapy at baseline | 1067.072 | 1063.670 | 5 | 5 | [793.315, 4398.717] | 7% |
| Baseline GAD-7 | 809.314 | 811.995 | 6 | 6 | [1125.229, 3980.259] | 0% |
| Suicide attempts (3 months) | 0.000 | 0.000 | 7 | 7 | [0.000, 0.000] | 0% |

*N = 502. Importance is expressed on the scale of the model objective function (residual sum of squares (continuous BSL-23 model)); values are comparable across moderators within this analysis but not across analyses. Cond. = conditional. Uncond. = unconditional. Imp. = importance. Freq. = frequency.*

**Supplementary Table 4. Variable importance for treatment response at T2 (exploratory).**

| Moderator | Cond. imp. | Uncond. imp. | Rank (cond.) | Rank (uncond.) | Conditional 95% CI | Root-split freq. |
| --- | --- | --- | --- | --- | --- | --- |
| Baseline BSL-23 | 9.065 | 9.184 | 1 | 1 | [5.405, 20.631] | 76% |
| Life-threatening events (3 months) | 3.168 | 3.076 | 2 | 2 | [1.292, 10.912] | 1% |
| Baseline PHQ-9 | 2.492 | 2.615 | 3 | 3 | [0.992, 12.419] | 1% |
| Medication at baseline | 1.581 | 1.493 | 4 | 4 | [0.224, 8.666] | 16% |
| Psychotherapy at baseline | 0.696 | 0.616 | 5 | 5 | [-0.130, 7.303] | 4% |
| Suicide attempts (3 months) | 0.000 | 0.000 | 6 | 6 | [0.000, 0.000] | 0% |
| Baseline GAD-7 | -0.293 | -0.215 | 7 | 7 | [-1.317, 6.294] | 1% |

*N = 502. Importance is expressed on the scale of the model objective function (log-likelihood (binary responder model)); values are comparable across moderators within this analysis but not across analyses. Cond. = conditional. Uncond. = unconditional. Imp. = importance. Freq. = frequency.*

**Supplementary Table 5. Variable importance for change in BSL-23 at T3 (exploratory).**

| Moderator | Cond. imp. | Uncond. imp. | Rank (cond.) | Rank (uncond.) | Conditional 95% CI | Root-split freq. |
| --- | --- | --- | --- | --- | --- | --- |
| Baseline BSL-23 | 3520.858 | 3472.090 | 1 | 1 | [3045.886, 15883.051] | 4% |
| Medication at baseline | 3286.987 | 3256.042 | 2 | 3 | [1922.850, 7562.597] | 61% |
| Baseline PHQ-9 | 3248.965 | 3278.700 | 3 | 2 | [2096.341, 9665.723] | 30% |
| Life-threatening events (3 months) | 1630.632 | 1525.050 | 4 | 4 | [924.754, 5244.143] | 3% |
| Psychotherapy at baseline | 1340.125 | 1342.519 | 5 | 5 | [1011.502, 5139.599] | 2% |
| Baseline GAD-7 | 719.479 | 691.010 | 6 | 6 | [1078.400, 4097.698] | 0% |
| Suicide attempts (3 months) | 0.000 | 0.000 | 7 | 7 | [0.000, 0.000] | 0% |

*N = 451. Importance is expressed on the scale of the model objective function (residual sum of squares (continuous BSL-23 model)); values are comparable across moderators within this analysis but not across analyses. Cond. = conditional. Uncond. = unconditional. Imp. = importance. Freq. = frequency.*

**Supplementary Table 6. Variable importance for treatment response at T3 (exploratory).**

| Moderator | Cond. imp. | Uncond. imp. | Rank (cond.) | Rank (uncond.) | Conditional 95% CI | Root-split freq. |
| --- | --- | --- | --- | --- | --- | --- |
| Life-threatening events (3 months) | 4.389 | 4.296 | 1 | 1 | [0.948, 13.835] | 1% |
| Baseline BSL-23 | 4.006 | 4.296 | 2 | 2 | [1.157, 13.207] | 83% |
| Medication at baseline | 1.928 | 1.854 | 3 | 3 | [0.155, 8.364] | 12% |
| Baseline PHQ-9 | 0.681 | 0.742 | 4 | 4 | [-0.317, 8.381] | 1% |
| Psychotherapy at baseline | 0.660 | 0.565 | 5 | 5 | [-1.056, 5.664] | 1% |
| Suicide attempts (3 months) | 0.000 | 0.000 | 6 | 6 | [0.000, 0.000] | 0% |
| Baseline GAD-7 | -1.615 | -1.479 | 7 | 7 | [-2.782, 5.499] | 2% |

*N = 451. Importance is expressed on the scale of the model objective function (log-likelihood (binary responder model)); values are comparable across moderators within this analysis but not across analyses. Cond. = conditional. Uncond. = unconditional. Imp. = importance. Freq. = frequency.*

### **S2. Partial-dependence profiles**

The following figures show partial-dependence (marginal CATE) profiles for the three most important moderators of each analysis: the model-based forest’s predicted treatment effect as a function of one moderator, averaged (marginalized) over the observed distribution of the remaining moderators. For the continuous BSL-23 outcome the vertical axis is the predicted change in BSL-23 (more negative = greater benefit of priovi); for treatment response it is the predicted risk difference (higher = greater benefit). Binary baseline indicators are displayed at their two levels (0 = absent, 1 = present). Profiles are descriptive and, like the subgroup analyses, should be read as hypothesis-generating.

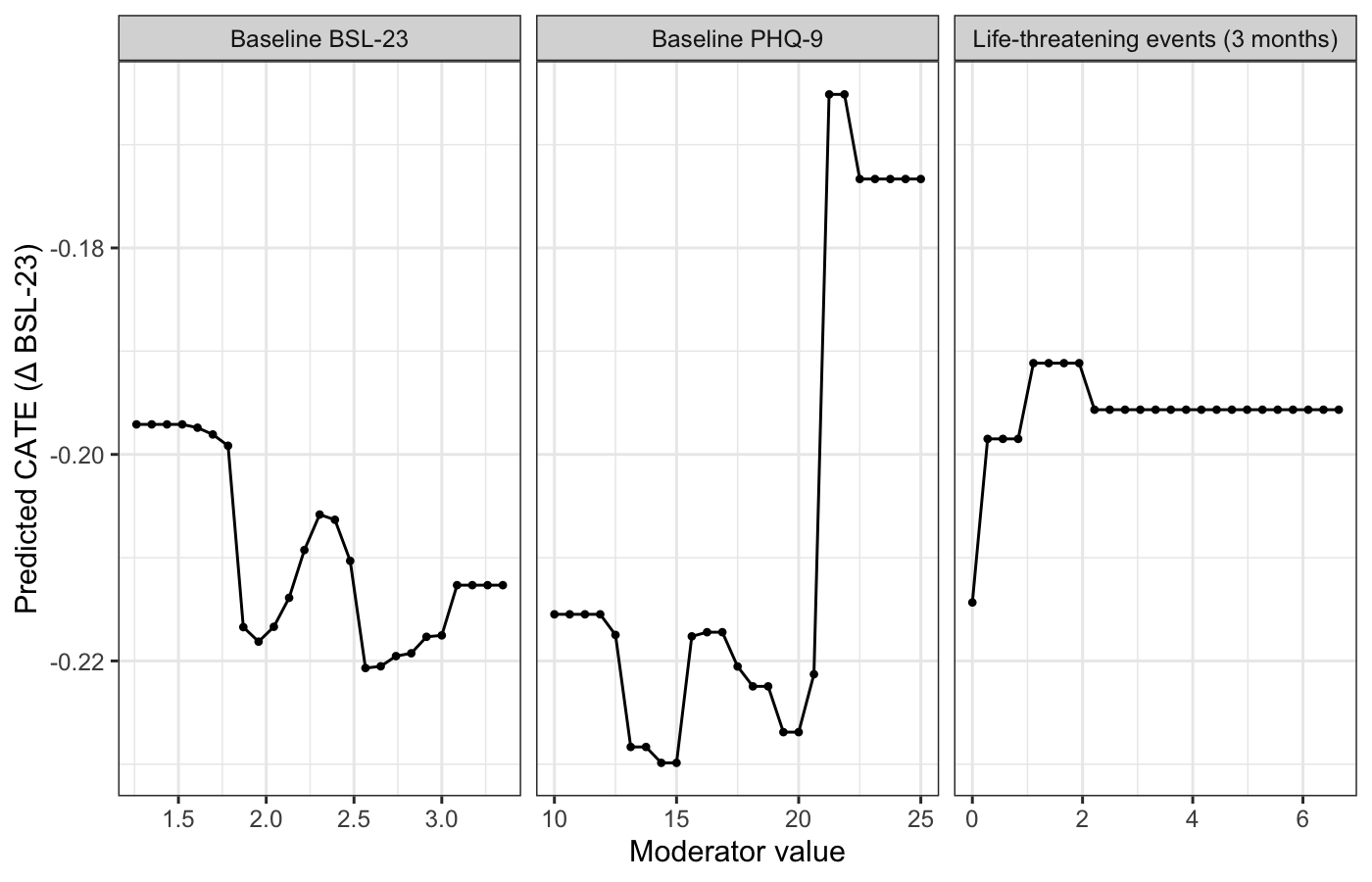

*Supplementary Fig. 1. Partial dependence of the estimated CATE on the three most important moderators (BSL-23 change at T1). Curves show the forest-predicted CATE while averaging over the other moderators (more negative = greater benefit of priovi). The axis is zoomed, so modest variation is visually exaggerated. Because PHQ-9 and baseline BSL-23 are correlated, the PHQ-9 profile, including the downturn at high scores, should not be interpreted as an isolated severity effect.*

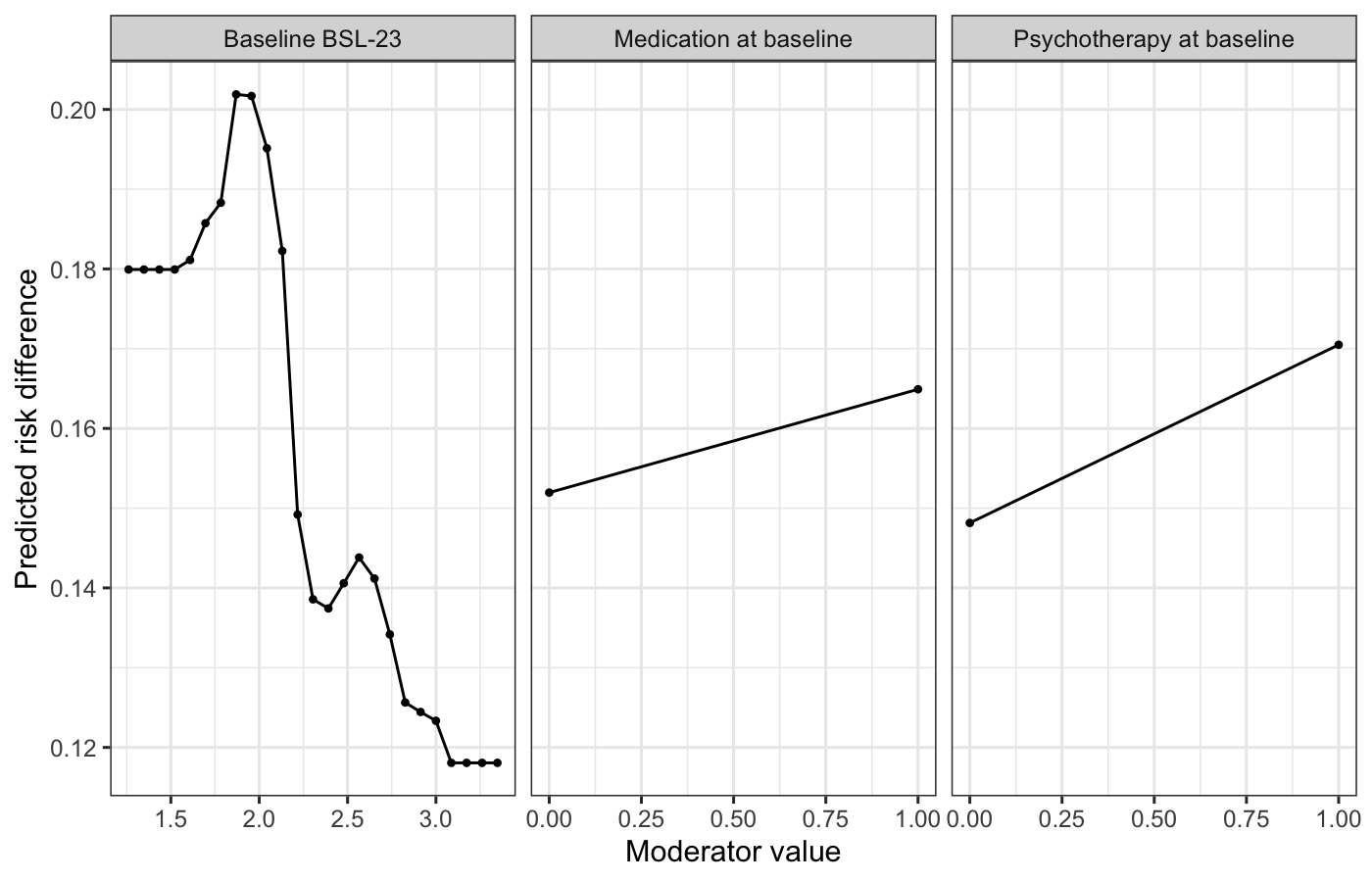

*Supplementary Fig. 2. Partial-dependence profiles for treatment response at T1.*

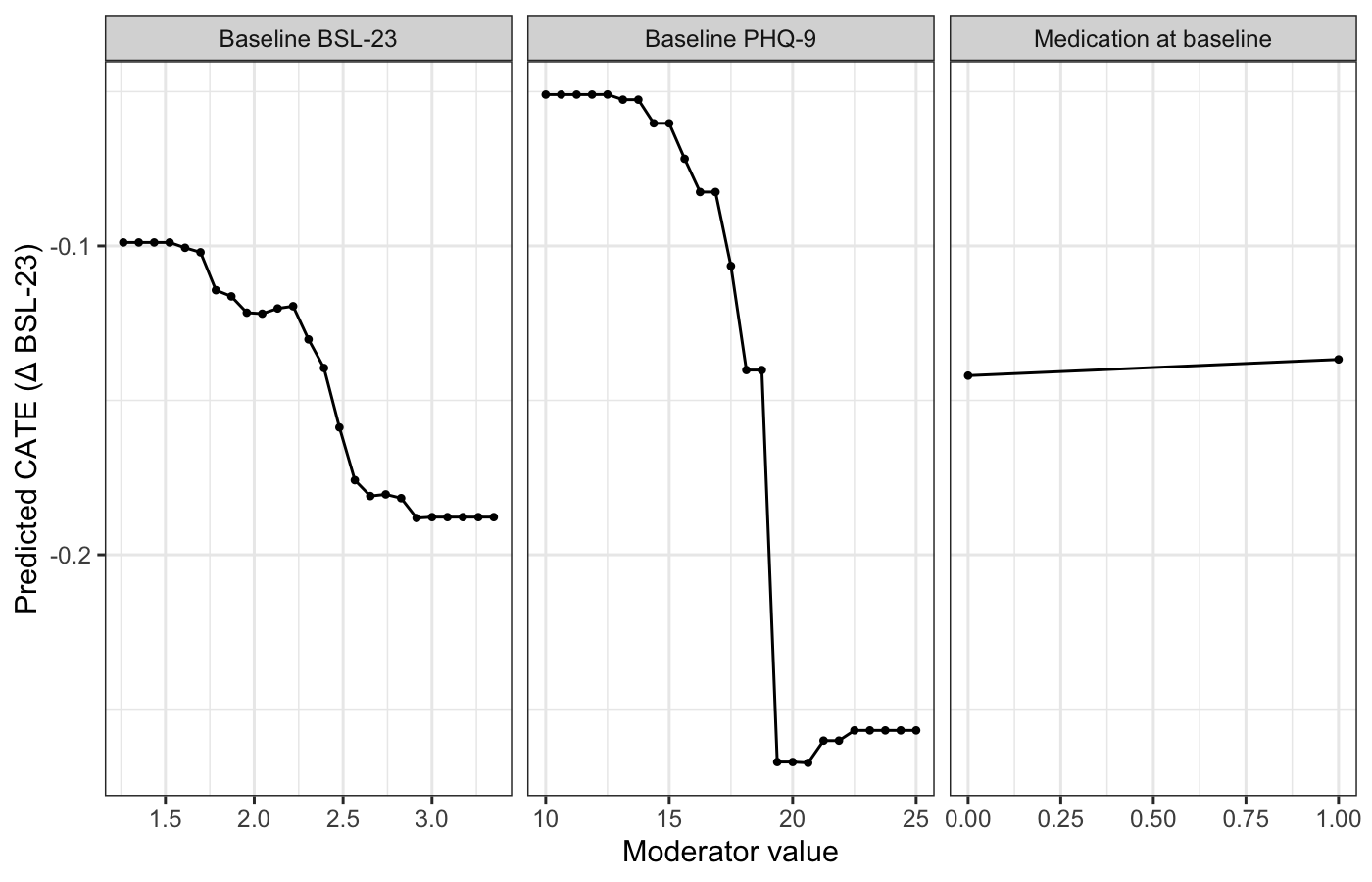

*Supplementary Fig. 3. Partial-dependence profiles for change in BSL-23 at T2 (exploratory).*

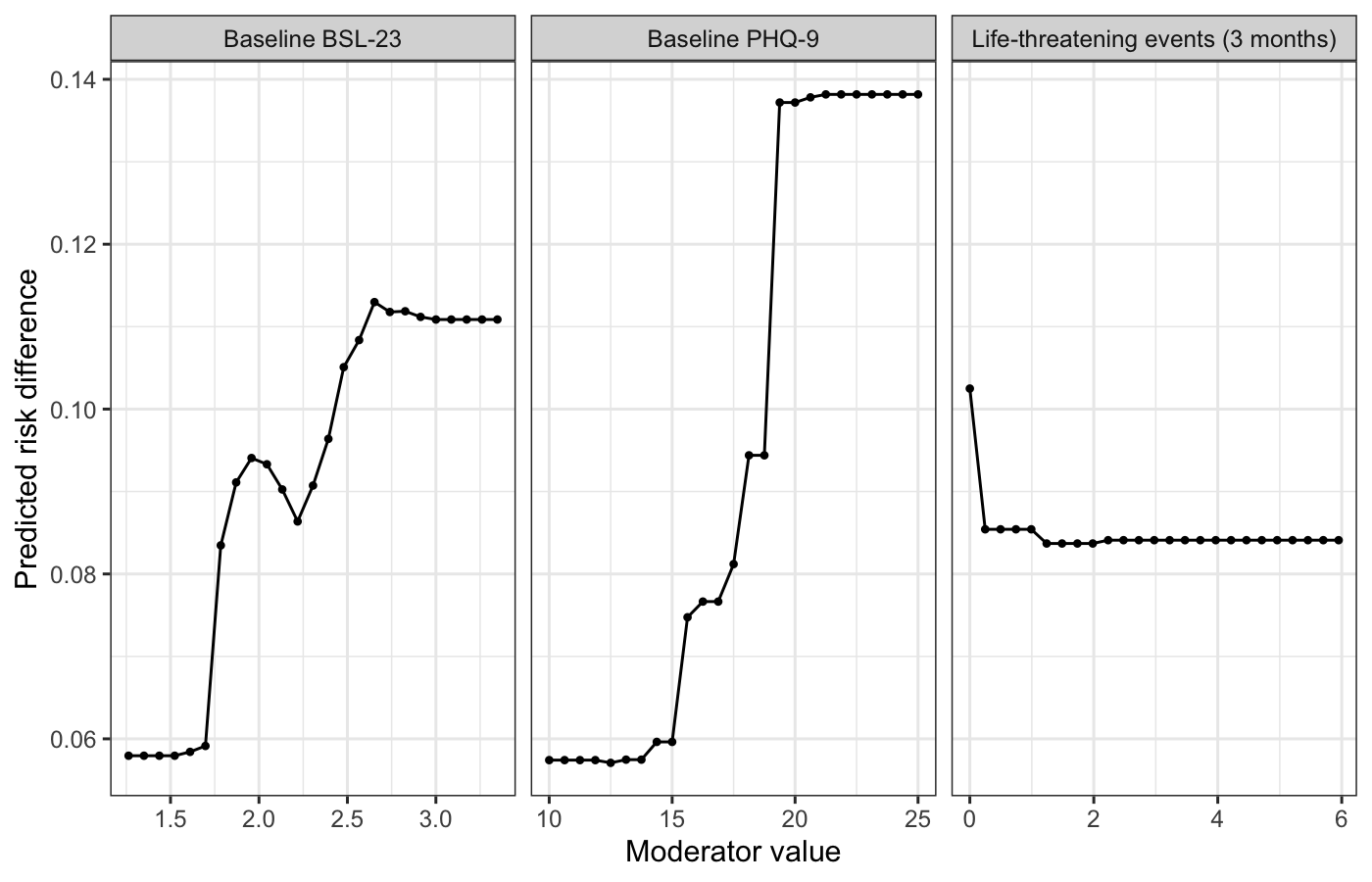

*Supplementary Fig. 4. Partial-dependence profiles for treatment response at T2 (exploratory).*

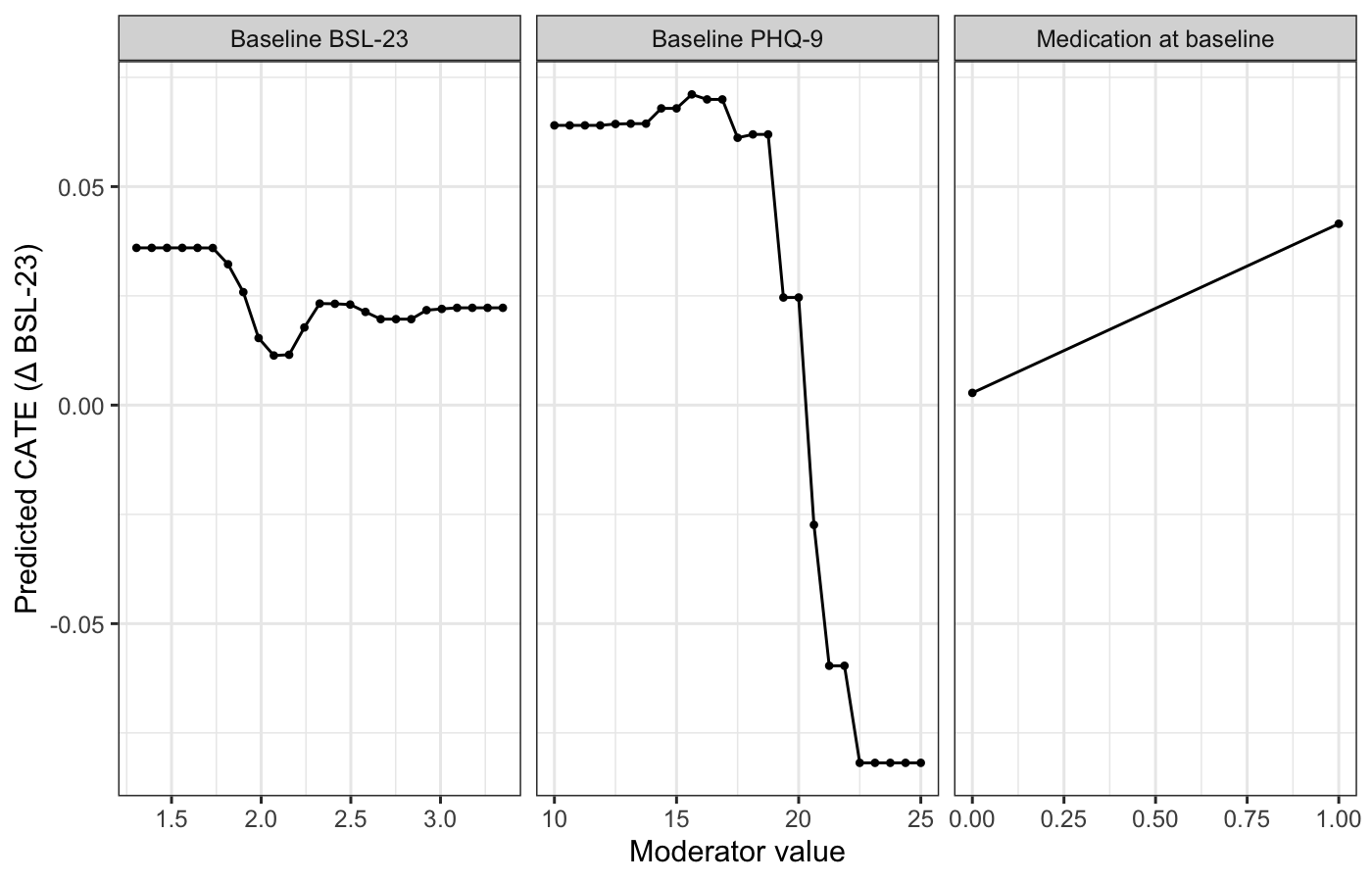

*Supplementary Fig. 5. Partial-dependence profiles for change in BSL-23 at T3 (exploratory).*

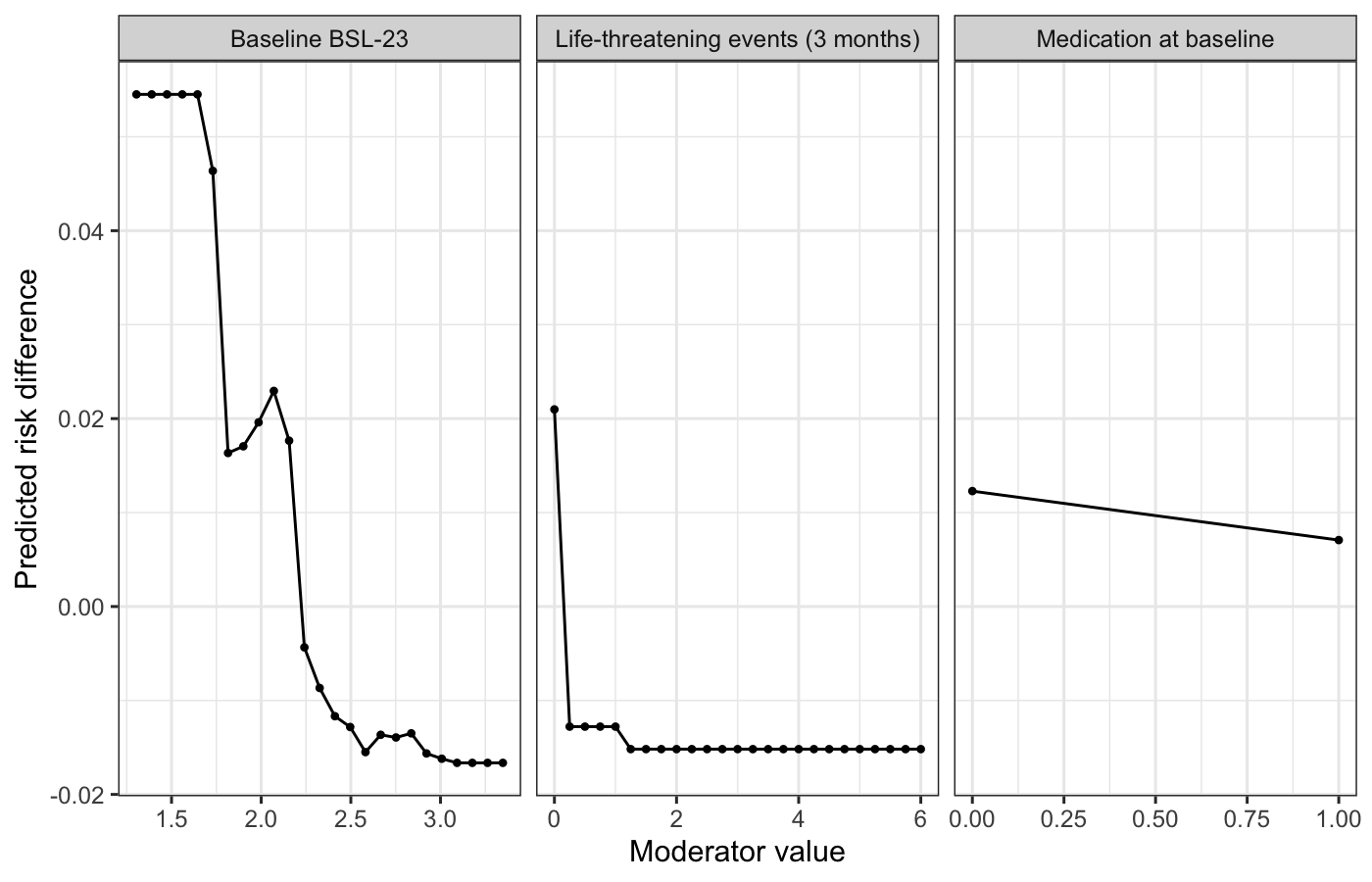

*Supplementary Fig. 6. Partial-dependence profiles for treatment response at T3 (exploratory).*

### **S3. Supplementary Note 1: sensitivity analysis (single stochastic imputation of the T1 outcome)**

The confirmatory analyses in the main text are complete-case: participants without an observed BSL-23 outcome at the primary endpoint (T1) are excluded. As a robustness check against differential dropout, we repeated the two T1 analyses after a single stochastic imputation of the follow-up BSL-23 outcomes, so that all randomized and baseline-complete participants (N = 578) contributed rather than only the T1 completers (N = 528).

Missing BSL-23 values at T1, T2 and T3 were imputed jointly by predictive mean matching (mice), fitted separately within each randomization arm so that arm-by-moderator relationships, and thus any treatment-effect heterogeneity, were preserved. Baseline moderators, which were complete by design, served only as predictors and were not imputed. A single completed dataset was drawn using the same random seed (1234) as the main analyses, and the binary responder outcome was derived from the imputed BSL-23 values using the identical reliable-change and severity-grade definition as in the primary analysis. All downstream steps (model-based forest, omnibus test, conditional variable importance, representative tree and stratified bootstrap) were identical to the complete-case pipeline and used the full specification of 500 trees with 1000 permutation-test and 1000 bootstrap replicates.

A single stochastic imputation was used rather than multiple imputation with Rubin pooling for computational reasons: the nested permutation omnibus test and the stratified bootstrap are computationally intensive, and repeating them across many imputed datasets would be prohibitive. The confidence intervals reported here are therefore conditional on the single imputed dataset and do not incorporate between-imputation uncertainty, so they are somewhat narrower than fully pooled intervals would be. This analysis probes the robustness of the substantive conclusions to differential dropout; the complete-case analysis remains the primary analysis ^2^.

Dropout at T1 was differential by arm (5.4 percent in the control arm versus 11.7 percent in the priovi arm). Of the 50 participants without an observed T1 outcome, 31 (62 percent) also had no observed T2 or T3 value, so for these participants the imputation draws essentially on baseline characteristics and arm; the remaining 19 contributed at least one later observation that informs the imputation.

Supplementary Table 7 compares the omnibus test, the most influential moderator and the representative-tree root split under the two approaches; Supplementary Tables 8 and 9 give the full conditional-versus-unconditional variable-importance comparison for the single-imputation analyses, and Supplementary Tables 10 and 11 give the corresponding subgroup treatment effects (to be read against Tables 3 and 5 of the main text). Broad agreement between the complete-case and single-imputation results indicates that the reported treatment-effect heterogeneity is not an artefact of the complete-case restriction.

**Supplementary Table 7. Complete-case versus single-imputation comparison at T1.**

| Analysis | N | Omnibus p | Top moderator (cond. VI) | Root split |
| --- | --- | --- | --- | --- |
| BSL-23 change, T1 (complete case) | 528 | 0.022 | Baseline BSL-23 | Baseline PHQ-9 at 23 |
| BSL-23 change, T1 (single imputation) | 578 | 0.028 | Baseline BSL-23 | Baseline PHQ-9 at 23 |
| Treatment response, T1 (complete case) | 528 | 0.007 | Baseline BSL-23 | Baseline BSL-23 at 1.83 |
| Treatment response, T1 (single imputation) | 578 | 0.017 | Baseline BSL-23 | Baseline BSL-23 at 2.13 |

*Omnibus p is the permutation heterogeneity-test p-value. “Top moderator” is the moderator with the highest conditional permutation importance. “Root split” is the moderator and cut-point selected at the root of the representative tree (BSL-23 on the 0-4 mean-item scale). Complete-case rows reproduce the main analysis; single-imputation rows use all N = 578 baseline-complete participants. Cond. VI = conditional variable importance.*

**Supplementary Table 8. Variable importance for change in BSL-23 at T1 (single-imputation sensitivity analysis).**

| Moderator | Cond. imp. | Uncond. imp. | Rank (cond.) | Rank (uncond.) | Conditional 95% CI | Root-split freq. |
| --- | --- | --- | --- | --- | --- | --- |
| Baseline BSL-23 | 10.218 | 9.954 | 1 | 1 | [8.074, 33.496] | 6% |
| Baseline PHQ-9 | 5.084 | 5.049 | 2 | 2 | [4.143, 15.493] | 72% |
| Life-threatening events (3 months) | 3.224 | 3.255 | 3 | 3 | [2.522, 11.100] | 9% |
| Baseline GAD-7 | 1.996 | 1.981 | 4 | 4 | [2.389, 8.922] | 4% |
| Medication at baseline | 1.910 | 1.822 | 5 | 5 | [1.868, 8.767] | 4% |
| Psychotherapy at baseline | 1.441 | 1.399 | 6 | 6 | [1.588, 7.867] | 4% |
| Suicide attempts (3 months) | 0.000 | 0.000 | 7 | 7 | [0.000, 0.000] | 1% |

*N = 578. Importance is expressed on the scale of the model objective function (residual sum of squares (continuous BSL-23 model)); values are comparable across moderators within this analysis but not across analyses. Cond. = conditional. Uncond. = unconditional. Imp. = importance. Freq. = frequency.*

**Supplementary Table 9. Variable importance for treatment response at T1 (single-imputation sensitivity analysis).**

| Moderator | Cond. imp. | Uncond. imp. | Rank (cond.) | Rank (uncond.) | Conditional 95% CI | Root-split freq. |
| --- | --- | --- | --- | --- | --- | --- |
| Baseline BSL-23 | 6.030 | 6.111 | 1 | 1 | [3.433, 18.923] | 77% |
| Baseline GAD-7 | 3.259 | 3.251 | 2 | 2 | [1.065, 12.121] | 5% |
| Medication at baseline | 1.955 | 1.909 | 3 | 3 | [0.944, 8.456] | 7% |
| Psychotherapy at baseline | 1.733 | 1.807 | 4 | 4 | [0.522, 8.967] | 11% |
| Baseline PHQ-9 | 1.593 | 1.603 | 5 | 5 | [0.915, 8.689] | 0% |
| Life-threatening events (3 months) | 1.458 | 1.561 | 6 | 6 | [0.795, 6.864] | 1% |
| Suicide attempts (3 months) | 0.000 | 0.000 | 7 | 7 | [0.000, 0.000] | 0% |

*N = 578. Importance is expressed on the scale of the model objective function (log-likelihood (binary responder model)); values are comparable across moderators within this analysis but not across analyses. Cond. = conditional. Uncond. = unconditional. Imp. = importance. Freq. = frequency.*

**Supplementary Table 10. Subgroup treatment effects, BSL-23 change at T1 (single-imputation sensitivity analysis).**

| Terminal node | CATE (Δ BSL-23) | 95% CI |
| --- | --- | --- |
| 2 | -0.205 | [-0.328, -0.077] |
| 3 | -0.360 | [-0.676, -0.079] |

**Supplementary Table 11. Subgroup treatment effects, treatment response at T1 (single-imputation sensitivity analysis).**

| Terminal node | Risk difference | 95% CI |
| --- | --- | --- |
| 2 | 0.246 | [0.127, 0.365] |
| 3 | 0.102 | [0.004, 0.204] |

### **S4. Consort 2025 Checklist**

| **Section / topic** | **No.** | **Checklist item** | **Reported on page no.** |
| --- | --- | --- | --- |
| **Title and abstract** |  |  |  |
| Title and abstract | 1a | Identification as a randomised trial | p. 1, Title: *Secondary Analysis of Treatment Effect Heterogeneity of priovi in Borderline Personality Disorder* (12 words; identifies the study as a secondary analysis). |
|  | 1b | Structured summary of the trial design, methods, results, and conclusions | p. 2, Abstract (unstructured, npj Digital Medicine; 146 words). Summarises design (secondary analysis of RCT; priovi + TAU vs TAU), primary endpoint T1, main heterogeneity results, and conclusion. Trial register and funding are not in the abstract (npj format); they appear in Methods / Acknowledgments. |
| **Open science** |  |  |  |
| Trial registration | 2 | Name of trial registry, identifying number (with URL) and date of registration | p. 14, Methods, Study design and participants: German Clinical Trials Register **DRKS00028888** (<https://drks.de/search/de/trial/DRKS00028888>; registered **27 April 2022**). Secondary-analysis preregistration: OSF <https://osf.io/sz72e/> (p. 14, Methods; also Code availability, p. 21). |
| Protocol and statistical analysis plan | 3 | Where the trial protocol and statistical analysis plan can be accessed | Parent trial: Assmann et al., 2025 (and trial registry/protocol as cited therein). Secondary-analysis plan: OSF preregistration <https://osf.io/sz72e/> (p. 14, Methods; p. 21, Code availability). Analysis code: OSF <https://osf.io/ker2t/> (p. 20, Statistics and reproducibility; p. 21, Code availability). |
| Data sharing | 4 | Where and how individual de-identified participant data, statistical code and other materials can be accessed | p. 21, Data availability: IPD not publicly available; available on reasonable request to J. P. Klein (methodologically sound proposal; up to 36 months after publication). p. 21, Code availability: full analysis code at OSF <https://osf.io/ker2t/>. |
| Funding and conflicts of interest | 5a | Sources of funding and other support, and role of funders | p. 21, Acknowledgments: secondary analysis received no specific funding; original trial funded by GAIA. |
|  | 5b | Financial and other conflicts of interest of the manuscript authors | p. 22, Competing interests (full author-level disclosures). |
| **Introduction** |  |  |  |
| Background and rationale | 6 | Scientific background and rationale | pp. 2–4, Introduction. |
| Objectives | 7 | Specific objectives related to benefits and harms | p. 4, Introduction (final paragraph): (1) treatment-effect heterogeneity; (2) most influential of seven candidate moderators; (3) whether higher baseline psychopathology is associated with larger absolute benefit; T2/T3 exploratory. Harms are not an objective of this secondary analysis (see Item 15 / 27). |
| **Methods** |  |  |  |
| Patient and public involvement | 8 | Details of patient or public involvement in the design, conduct and reporting of the trial | p. 20, Methods, Patient and public involvement: patients and the public were not involved in design, conduct, analysis, or reporting of this secondary analysis; in the parent trial, individuals with BPD were involved in developing priovi but not in trial design (Assmann et al., 2025). |
| Trial design | 9 | Description of trial design (type, allocation ratio, framework) | p. 14, Methods, Study design and participants: pragmatic, rater-blinded, two-arm parallel-group RCT; 580 randomised (302 priovi + TAU, 278 TAU). Framework/allocation details of the parent trial: Assmann et al., 2025. |
| Changes to trial protocol | 10 | Important changes after commencement, including non-prespecified outcomes/analyses | p. 20, Methods, Deviations from the preregistration (exclusion of 2 participants with incomplete baseline moderators; N = 528 vs preregistered 530). Analytic pipeline otherwise as preregistered. Parent-trial amendments: Assmann et al., 2025. |
| Trial setting | 11 | Settings and locations where the trial was conducted | p. 14, Methods: randomised in Germany. Detailed site/setting description: Assmann et al., 2025. |
| Eligibility criteria | 12a | Eligibility criteria for participants | p. 14, Methods, Study design and participants (SCID-5-PD BPD; BSL-23 mean ≥ 1.07; exclusions for substance use / psychotic disorder). Full criteria: Assmann et al., 2025. |
|  | 12b | Eligibility criteria for sites and individuals delivering interventions (if applicable) | Assmann et al., 2025 (digital self-guided intervention; site/delivery criteria of parent trial). |
| Intervention and comparator | 13 | Intervention and comparator with sufficient details to allow replication | p. 15, Methods, Treatment condition (priovi + TAU vs TAU). Full intervention description and materials: Assmann et al., 2025. |
| Outcomes | 14 | Prespecified primary and secondary outcomes (variable, metric, aggregation, time point) | pp. 15–16, Methods, Measures: continuous BSL-23 mean-item change; binary clinically relevant response (RCI + severity-grade shift); confirmatory time point T1 (3 months); T2/T3 exploratory (p. 14). Parent-trial primary endpoint context: Assmann et al., 2025. |
| Harms | 15 | How harms were defined and assessed | Not an objective of this secondary heterogeneity analysis. Parent-trial harms assessment and reporting: Assmann et al., 2025. |
| Sample size | 16a | How sample size was determined | Parent-trial sample size: Assmann et al., 2025. Present paper uses the randomised EPADIP-BPD sample with complete-case follow-up; Discussion notes power for heterogeneity (about d ≥ 0.35) (p. 11). |
|  | 16b | Interim analyses and stopping guidelines | Assmann et al., 2025 (parent trial). Not applicable to this secondary analysis. |
| Randomisation: sequence generation | 17a | Who generated the random allocation sequence and method used | Assmann et al., 2025. |
|  | 17b | Type of randomisation and any restriction (stratification, blocking) | Assmann et al., 2025. |
| Allocation concealment mechanism | 18 | Mechanism used to implement the allocation sequence | Assmann et al., 2025. |
| Implementation | 19 | Who enrolled participants and who assigned participants to interventions | Assmann et al., 2025. |
| Blinding | 20a | Who was blinded after assignment (participants, care providers, outcome assessors) | p. 14, Methods: rater-blinded parent trial; Assmann et al., 2025 for full blinding details. |
|  | 20b | If blinded, how blinding was achieved and similarity of interventions | Assmann et al., 2025. |
| Statistical methods | 21a | Statistical methods used to compare groups for primary and secondary outcomes, including harms | pp. 17–20, Methods, Statistical analysis (MOB-RP / model-based forests; omnibus test; model assumptions and inference; variable importance; representative tree; CATEs; bootstrap CIs; calibration; c-for-benefit; multiplicity; statistics and reproducibility). Harms analyses: Assmann et al., 2025. |
|  | 21b | Definition of who is included in each analysis, and in which group | p. 16, Methods, Missing data and analysis samples; Results sample sizes by time point (pp. 4–8). Complete-case by follow-up; groups = randomised arms (control / intervention). |
|  | 21c | How missing data were handled | p. 16, Methods, Missing data and analysis samples (complete-case primary). pp. 17–18, Methods, Statistical analysis + Supplementary Information: single stochastic imputation sensitivity analysis at T1. |
|  | 21d | Methods for any additional analyses (subgroup / sensitivity), distinguishing prespecified from post hoc | pp. 17–20, Methods, Statistical analysis and Multiplicity: confirmatory T1 analyses and exploratory T2/T3 as preregistered; single-imputation sensitivity in Supplementary Information. p. 20, Deviations from the preregistration. |
| **Results** |  |  |  |
| Participant flow | 22a | For each group, numbers randomly assigned, received intended intervention, and analysed for the primary outcome | Randomised N and arm sizes: p. 14, Methods (580; 302 / 278). Analysis N at T1: p. 4, Results / Sample characteristics; p. 25, Table 1 (528; 263 / 265). Full CONSORT flow (screened, excluded, received intervention): Assmann et al., 2025. No new flow diagram in this secondary paper. |
|  | 22b | For each group, losses and exclusions after randomisation, with reasons | p. 16, Methods: complete-case restriction; differential dropout by arm; p. 20, exclusion of 2 participants with incomplete baseline moderators (Deviations). Detailed attrition reasons: Assmann et al., 2025. |
| Recruitment | 23a | Dates defining periods of recruitment and follow-up | Assmann et al., 2025. Follow-up schedule: p. 14, Methods (T0, T1 3 months, T2 6 months, T3 12 months). |
|  | 23b | Why the trial ended or was stopped (if relevant) | Assmann et al., 2025 (if applicable); otherwise N/A. |
| Intervention and comparator delivery | 24a | Intervention and comparator as actually administered | Assmann et al., 2025. |
|  | 24b | Concomitant care received during the trial for each group | Assmann et al., 2025 (TAU context). |
| Baseline data | 25 | Table of baseline demographic and clinical characteristics for each group | p. 25, Table 1 (T1 confirmatory complete-case sample by arm); introduced p. 4, Results. Full randomised-sample baseline table: Assmann et al., 2025. |
| Numbers analysed, outcomes and estimation | 26 | For each primary and secondary outcome, by group: n analysed; n with available data; result per group; effect size and precision; for binary outcomes absolute and relative effects | pp. 5–8, Results (T1 BSL-23 and responder; exploratory T2/T3). Subgroup CATEs with 95% CIs: p. 25-26, Tables 3 and 5; Figures 1 and 3 (pp. 23, 25). Responder rates by arm at T1 (41% vs 57%; p. 6). Average treatment effects of the parent trial: Assmann et al., 2025. |
| Harms | 27 | All harms or unintended events in each group | Not reported in this secondary analysis. Assmann et al., 2025. |
| Ancillary analyses | 28 | Any other analyses, including subgroup and sensitivity analyses, distinguishing prespecified from post hoc | pp. 5–8, Results: preregistered confirmatory heterogeneity analyses at T1; exploratory T2/T3; Supplementary Information (conditional vs unconditional importance; partial-dependence profiles; single-imputation sensitivity). All framed as secondary/ancillary to Assmann et al., 2025. |
| **Discussion** |  |  |  |
| Interpretation | 29 | Interpretation consistent with results, balancing benefits and harms, considering other evidence | pp. 9–13, Discussion. Harms balance refers to parent-trial evidence / Assmann et al., 2025. |
| Limitations | 30 | Trial limitations (bias, imprecision, generalisability, multiplicity) | pp. 11–12, Discussion (complete-case / MAR, power for heterogeneity, prognostic vs moderating splits, small high-benefit subgroup, weak validation of predicted treatment effects, attenuation at 12 months, rare suicide-attempt moderator, single-trial generalisability). |
